# Developing a core outcome set for interventions in people with mild cognitive impairment: study protocol

**DOI:** 10.1101/2024.07.05.24309989

**Authors:** Victoria Grace Gabb, Sam Harding, Angus McNair, Julie Clayton, Winsome Barrett-Muir, Alan Richardson, Natalie Woodward, Sophie Alderman, Jemima Dooley, Joseph Webb, Sarah Rudd, Elizabeth Coulthard, Nicholas Turner

## Abstract

**Introduction:** There is no standardised national guidance on clinical management for people living with mild cognitive impairment (MCI) and therapeutic interventions are limited. Understanding what outcomes are important and meaningful to people living with MCI and developing a core outcome set for research and clinical practice will improve the impact of clinical research and contribute towards developing effective care pathways for MCI. This study aims to develop a core outcome set (COS) for adults living with MCI intended for use in interventional and clinical settings.

**Methods and analysis:** The COS will be developed using a five-stage study design: (1) systematic literature search; (2) qualitative interviews; (3) evidence synthesis from stage 1 and 2; (4) two-round Delphi survey; (5) consensus meetings. First, we will conduct an umbrella review of existing MCI interventional studies and extract a list of outcomes. Qualitative interviews will be held with key stakeholders including individuals living with MCI, friends and family, and relevant professionals to identify further outcomes considered important. Outcomes from the review and interviews will be synthesised into a “long list” of outcomes for potential inclusion in the COS. Two rounds of a Delphi surveys followed by a consensus meeting will be used to reach stakeholder consensus on which outcomes should be included in the final COS.

**Ethics and dissemination:** We have received ethical approval from London - Queen Square Research Ethics Committee (23/PR/1580). Patient and public involvement and engagement are central to developing the COS. The results will be disseminated via conferences, peer-reviewed publications, briefing notes to key agencies, to the public via social media and blog posts, and directly to stakeholders who participate in the project.

**Trial registration number:** Core Outcome Measures in Effectiveness Trials (COMET) Initiative 2117; PROSPERO registration: CRD42023452514.

**Strengths and limitations:**

- To our knowledge, this will be the first core outcome set developed to evaluate interventions for people with mild cognitive impairment (MCI).
- The study will use a multi-stage process including an umbrella review, qualitative interviews, Delphi surveys, and a consensus meeting to incorporate multidisciplinary stakeholder perspectives including those living with MCI and individuals who know, care for, work with, or provide services for people with MCI.
- Development and adoption of this COS is expected to enhance the value of research into MCI and clinical practice through encouraging transparent reporting of agreed meaningful outcomes to stakeholders.
- A limitation of this study is that recruitment sites will be based in England only. However, we will aim to recruit international stakeholders to participate in the study and the umbrella review will include international literature. Efforts will be made to include individuals at each qualitative stage with diverse experience, backgrounds, and demographics.
- There are many possible outcomes for inclusion in the COS and it may be difficult to reach consensus across stakeholders.

## Introduction

### Background and objectives

Mild cognitive impairment (MCI) is a common clinical syndrome describing cognitive symptoms and objective cognitive impairment on neurocognitive testing without significant impairment on daily functioning^1^. MCI is the most common diagnosis after dementia in memory services in England and Wales ^2^, and recent worldwide prevalence estimates suggest that MCI affects approximately 15% of people living in the community over the age of 50^3^, though numbers of people seeking diagnosis in the early stages of memory or cognitive impairment may increase as disease-modifying treatments become available. The risk of developing MCI increases with age and decreases with education level^3^.

Often described as a stage between normal age-related cognitive decline and dementia, MCI is a risk factor for dementia, with estimated annual rates of progression from MCI to dementia at 8-16%^1^. Factors predicting more rapid progression to dementia include being diagnosed with amnestic or multi-domain MCI, having biomarkers of Alzheimer’s disease (AD) such as higher amyloid burden^4^, apolipoprotein E (APOE) ε4 carrier status, and co-morbid frailty and depression^1^.

There are currently no standardised NICE guidelines for the management of MCI, and international guidelines and recommendations are inconsistent^5^. Clinical pathways vary regionally in the UK, but typically people diagnosed with MCI are informed of an increased risk of developing dementia and are either discharged from memory services or may be monitored for progression to dementia, without tailored symptomatic therapy^6^.

There is some evidence that exercise and cognitive training can improve cognitive outcomes in MCI^7^, and there has been recent progress in disease-modifying anti-amyloid drugs for AD at the MCI stage, though debate continues on how to define clinically meaningful outcomes^8^. The heterogeneity of outcomes used in MCI research prevents comparisons across interventions and inhibits meaningful synthesis in systematic reviews and meta-analyses, limiting clinical impact^9^. Lack of stakeholder and patient input into outcomes selected for trials also means selected primary outcomes may not be the most meaningful outcomes to people with MCI^9,10^. Individuals with MCI want to be actively involved in the management and decision-making regarding their healthcare^11^. Defining the effectiveness of interventions in terms of what outcomes are most important to key stakeholders–individuals living with MCI and the people who know, care for, work with, and provide services to them–through developing an agreed core outcome set (COS) will help to establish the most effective interventions for people with MCI.

A COS is a standardised minimum set of outcomes which researchers are recommended to measure in trials for a specific health condition^12^. Implementing COS helps with evidence synthesis and more confident recommendations, and helps to ensure outcomes that are important to the people most affected by the health condition are evaluated and published^12^. Though most often considered in regard to interventional and clinical trials, COS can also be useful in standardising the collection of clinical data, allowing for analysis of change over time, and are also utilised by health technology assessments^13^. Several COS have been established in people with dementia^14,15^. However, what matters to those with MCI may vary to those with a diagnosis of dementia. Not all individuals with MCI progress to dementia and the rate of cognitive decline can vary substantially^16^. MCI also often affects people of working age or in early retirement and clinical symptoms differ significantly from people with dementia. Accordingly, the NICE guidelines for dementia currently exclude patients with an MCI diagnosis. There is currently no COS for people living with MCI.

In this study, we will establish a COS of importance to people with lived experience of MCI for use in MCI research and clinical practice, to help optimise research and standardise care.

Specifically, we aim to:

1. Identify what outcomes have been used in MCI studies to date by conducting a systematic umbrella review.

2. Elicit stakeholder views on what outcomes they consider important through conducting qualitative interviews.

3. Integrate outcomes identified through the review and interviews and asking stakeholders to rate their importance and reach consensus on the final COS recommendations.

### Scope

The COS will be suitable for adults living in the community diagnosed with any type of MCI. The COS will be most suitable for interventional (pharmacological and non-pharmacological) research and may also be useful in clinical practice.

## Methods

This study is registered with the Core Outcome Measures in Effectiveness Trials (COMET) database (Reference #2117). The study has been designed in accordance with the COMET handbook^17^ and Core Outcome Set-STAndards for Development (COS-STAD) recommendations^18^. The protocol is reported in line with guidance from the Core Outcome Set-STAndardised Protocol (COS-STAP)^19^ items.

The study will consist of five stages (Figure 1):

1. Systematic umbrella review of reported outcomes for adults with MCI;
2. Individual interviews with stakeholders (e.g., individuals with MCI, partners, healthcare professionals);
3. Evidence synthesis from Stage 1 and Stage 2 to create a list of possible outcomes to include in the COS;
4. Two rounds of Delphi surveys among stakeholders to identify further outcomes and prioritise outcomes;
5. Consensus meeting(s) to agree on the final COS recommendations.

**Figure 1.**
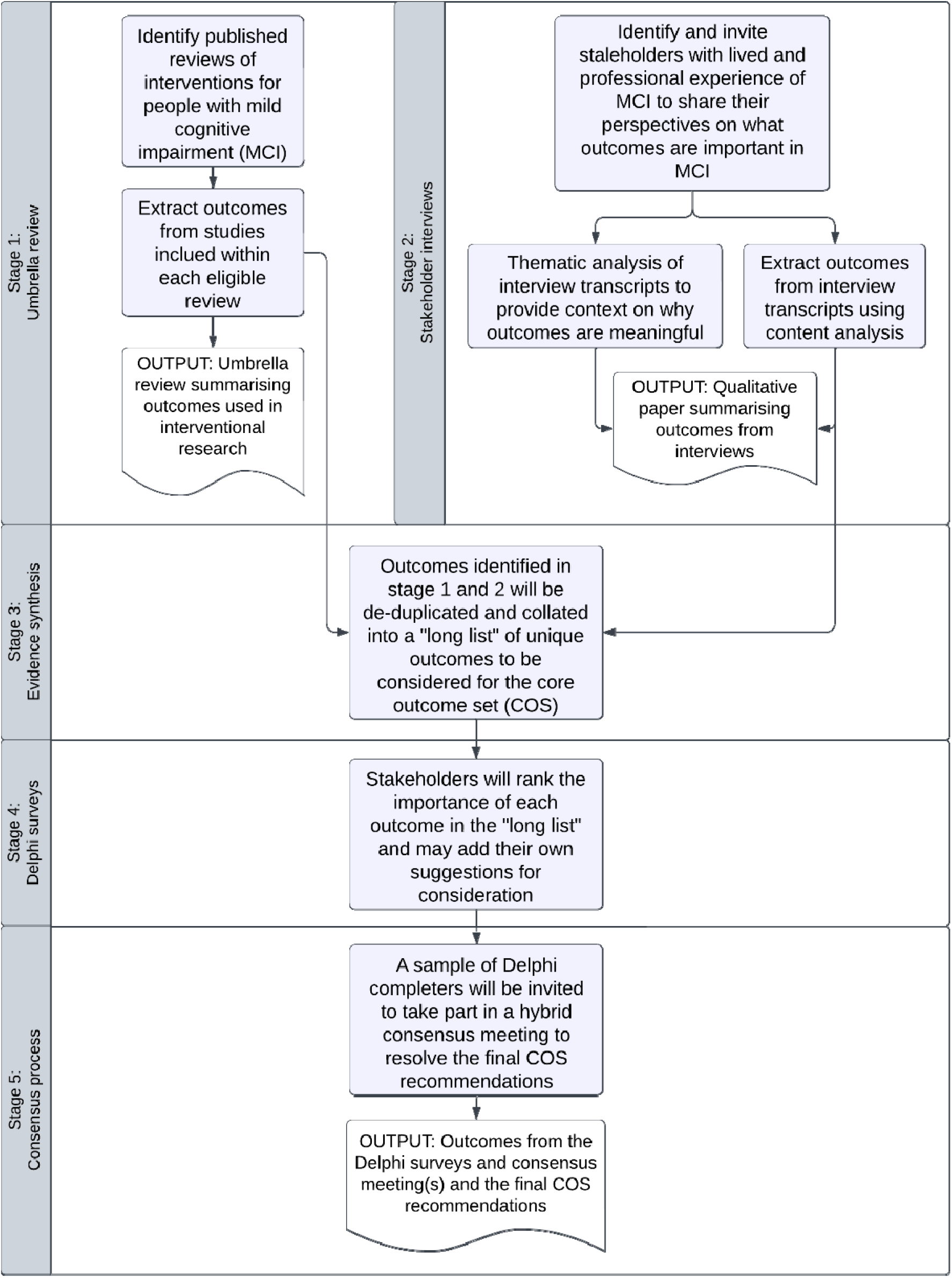
Study flowchart to demonstrate development of the mild cognitive impairment (MCI) core outcome set (COS) and anticipated outputs.

### Stakeholders

We have identified three stakeholder groups who have lived and/or professional experience of MCI, who will be involved in the COS development process:

1. Individuals diagnosed with MCI in the last 12 months (to minimise likelihood of progression to dementia since diagnosis);
2. Partners, family members, friends, and/or carers of people with MCI;
3. Professionals who work with or provide services for people with MCI (e.g., health and social care professionals, researchers, policymakers, people from community or faith groups).

Stakeholders will be invited to take part in interviews to identify outcomes and in Delphi surveys and consensus meetings to reach consensus on the COS.

### Patient and public involvement

Patient and public involvement (PPI) has been integral to study design and will continue to guide the study throughout. We have two PPI co-applicants who have experience with research, MCI, and dementia and whom form part of the study management group alongside the research team. During study development, regular PPI meetings with individuals with lived experience of MCI and dementia have helped to design and improve the protocol, participant-facing documents, and interview topic guides. During the study, PPI will be sought at regular intervals depending on the interests and expertise of the group (e.g., guiding how we disseminate findings and engage with participants, advising on wording of the outcomes in the Delphi survey).

### Information sources

#### Stage 1: Systematic umbrella review

Given the extensive literature conducted into MCI, we will conduct a systematic umbrella review of scoping and systematic reviews of interventional studies in MCI.

##### Eligibility criteria for inclusion in the review

For a review to be included, it must be either a systematic or scoping review published in English and over 70% of the research studies within the review must meet the following criteria: an interventional study involving community-dwelling adults diagnosed with MCI based on clinical diagnosis and/or in line with standardised MCI criteria (e.g., Petersen criteria^20^). Where an individual study involves a mixed cohort (e.g., participants with MCI and participants with dementia), the results must be available separately for the participants with MCI to be eligible.

##### Search strategy

An initial scoping search was undertaken using Medline (Ovid) to identify relevant articles on the topic. The text words contained in the titles and abstracts, and the subject headings for these articles were used to develop a full search strategy. The final search strategy was developed by an experienced medical librarian in collaboration with the research team and will be adapted for each included database (Embase (Ovid), CINAHL (EbscoHost), PsycINFO (Proquest), Medline (Ovid), COMET) and the PROSPERO register.

##### Study selection

Following the searches, all identified citations will be collated, uploaded to Endnote, and de-duplicated. De-duplicated citations will be uploaded to Rayyan^21^. Titles and abstracts will be screened by two or more independent reviewers for assessment against the inclusion criteria. Potentially relevant sources will be retrieved in full and uploaded to Rayyan. The full text of selected citations will be assessed in detail against the inclusion criteria by two or more independent reviewers. Reasons for exclusion at full-text screening will be recorded and reported in the final review. Any disagreements that arise between the reviewers at each stage of the selection process will be resolved through discussion, or with an additional reviewer. The results of the search and the study inclusion process will be reported in full in the final umbrella review.

##### Data extraction

We will retrieve the full text of the original studies included in eligible reviews to ensure accurate and detailed reporting of the outcomes collected. Duplicates will be removed before data extraction and where multiple records identifying different outcomes are identified for a single study, these will be collated into a single line in the data extraction form to prevent duplication in the results. The data extracted will include details about the participants (e.g., criteria used to define MCI, age, sex), study methods (e.g., study design, type of intervention), context (e.g., geographic region), specific outcomes reported, and how outcomes were operationalised (e.g., instrument or definition used, timeframe). The verbatim wording of outcomes (outcome definitions and/or measurement instruments) will be extracted from the source manuscript, before being coded using Dodd’s taxonomy^22^ into ‘outcomes’, ‘domains’, and ‘core areas’. The relevance of the taxonomy will be reviewed by the study management group and PPI contributors and may be amended if required^23^.Data extraction will be completed using a data extraction form. The draft data extraction tool will be modified and revised as necessary during the process of extracting data from each included evidence source and modifications will be detailed in the umbrella review and presented alongside the final data extraction form.

##### Data analysis and presentation

A flowchart based on the Preferred Reporting Items for Systematic reviews and Meta-Analyses (PRISMA) flowchart ^24^ will be used to chart records retained or removed at each stage of the umbrella review: number of records identified through database searching, records remaining following de-duplication, and exclusion of studies at various stages of screening (title, abstract, full text), to contextualise the final number of records included in the review. Study characteristics (e.g., year, intervention, outcomes) will be summarised and presented tabularly. Specific outcome measures will be tabulated and classified into domains (e.g., quality of life, cognition) and core areas (e.g., physiological/clinical, life impact resource use) and presented alongside key study characteristics. We will report on the number of unique specific outcome measures used and the frequency of each specific outcome measure, domain, and core area across all studies.

#### Stage 2: Qualitative interviews

Outcomes identified in the published literature may represent outcomes important to researchers and/or Sponsors and funding agencies. Interviews will be conducted to capture outcomes considered important to individuals with experience of MCI.

##### Participant recruitment

Participants with lived or professional experience of MCI will be invited to take part in the interviews, including:

1. Individuals diagnosed with MCI in the last 12 months (to minimise likelihood of progression to dementia since diagnosis);
2. Partners, family members, friends, and/or carers of people with MCI;
3. Professionals who work with or provide services for people with MCI (e.g., health and social care professionals, researchers, policymakers, people from community or faith groups).

The concept of information power will be used to determine sample size^25^, however we anticipate conducting 45 to 50 interviews in total. This includes approximately 20-25 interviews with those living with MCI, 15 interviews with friends, relatives, partners, and/or carers, and 10-15 interviews with other/professional stakeholders (e.g., health and social care professionals, researchers, and policymakers). Recruitment will primarily be via NHS research sites in England, engagement with community groups, voluntary/charitable organisations, and links with relevant research and professional networks. Recruitment may also be supported by social media advertisements if required.

##### Study design

Interviews will be conducted one-to-one with a researcher, either virtually or in person. Participants will receive information sheets and can ask any questions in advance of the meeting. Informed consent will be received either via e-Consent or paper consent forms. The interviews will last approximately 30-45 minutes. A topic guide has been co-developed by qualitative experts and our PPI group. Sampling will be purposive and aim to provide a broad representation of stakeholders and characteristics. As such, basic demographics data (e.g., age, sex, ethnicity) will be requested from all participants. Key clinical characteristics (e.g., date of diagnosis, likely aetiology of MCI, results of biomarker testing if available) will be recorded from participants with MCI to help characterise the cohort.

##### Data analysis

Virtual meetings will be recorded using the virtual meeting platform. In-person meetings will be recorded via an encrypted recording device. Recordings will be transcribed and analysed via content analysis to identify all possible outcomes of importance to stakeholders, and a thematic analysis will explore why these outcomes are meaningful.

#### Stage 3: Evidence synthesis

Findings from the umbrella review and qualitative interviews will be synthesised into a “long-list” of outcomes within domains. They will be consolidated and de-duplicated using a structured approach until no further outcomes are identified. Each verbatim outcome definition will be categorised to an outcome name, and each outcome name mapped to an outcome domain using an interactive focus group approach. The process will also be guided by the thematic analysis of the interviews in stage 2.

A plain English statement will be co-produced with PPI contributors to describe each outcome of the “long-list”, avoiding the use of jargon and technical terms (medical terminology may be included in parentheses). The study management group will review these statements for interpretability and make revisions and/or consult with the PPI group if required.

#### Stage 4: Delphi surveys and consensus meetings

Stakeholders will be invited to two rounds of anonymous Delphi surveys to generate consensus about the outcome domains of importance to key stakeholders.

##### Participant recruitment

Participants from the Stage 2 qualitative interviews will be invited to participant in the Delphi surveys. Additional participants will be recruited using the same recruitment methods as in Stage 2 to reach a target total of 124 participants, which is the median number of participants in Delphi surveys for COS development studies^26^.

##### Study design: Delphi surveys

Prior to the surveys, participants will be sent information explaining the purpose and context of the survey and plain language summaries including descriptions of what is meant by an outcome, a COS, and a Delphi survey. The study management group will develop accessible lay definitions of each outcome, including medical terms in parentheses where this may be helpful (e.g., for professionals).

Under the Delphi technique, experts are asked their opinions in a series of “rounds”, with the opportunity to see the anonymous results of previous rounds to enable reflection and repositioning of opinions to reach a consensus^27^. Delphi surveys are increasingly used to determine which outcomes should be included in a COS^28^.

Participants will be provided with the “long-list” of outcomes from Stage 3, with accessible (lay and medical) descriptions and asked to rate on a nine-point Likert scale how important it is to include each outcome in the COS (1 = “not essential” to 9 = “absolutely essential”). Two rounds will be conducted, and participants can complete the Delphi surveys either online or via paper surveys. In round one, participants will rate each outcome and will have the option to add additional outcomes. Between round one and two, the study management group will develop accessible statements for any outcomes added during round one. For round two, participants will be reminded of their round one rankings, and how other participants on average in each of the three stakeholder groups ranked each outcome. They will have the opportunity to review and change their responses if they would like to.

In case of attrition between the first and second Delphi survey, a sensitivity analysis will be used to assess whether the reduced sample affects conclusions drawn.

##### Study design: Consensus process

The percentage agreement of outcome importance in round two of the Delphi surveys will be used to determine consensus:

- Consensus in: ≥70% scored statement ≥6, and less than 10% scored statement ≤2.
- Consensus out: ≥70% scored statement ≤4, and less than 10% scored statement ≥8.

A consensus meeting will be held to discuss outcomes not meeting pre-specified Delphi criteria for inclusion/exclusion (defined above), and any outcomes where there is disagreement (i.e., Delphi consensus ‘in’ by one stakeholder group and ‘out’ by another). In round two of the Delphi survey, a final question will be asked about willingness to participate in a consensus meeting.

Participants in the consensus meeting will be sampled from the Delphi completers. The consensus meeting will be held as a hybrid (virtual and in-person) meeting, and depending on the needs (e.g., availability, fatigue) and preferences of the group, this meeting may be split into shorter meetings held within a week of each other.

The final output from the study will be a set of core outcomes of importance to people with MCI, as agreed by stakeholders.

## Discussion

This study protocol presents the methodology for the development of a COS for MCI research. Developing a COS is an important first step in improving outcome measurement and will be applicable for observational and interventional studies for adults with MCI. The COS may also support more comprehensive clinical reporting. Once the COS is established, further research will be needed to identify the optimal measurement methods or instruments available for each core outcome.

### Strengths

There is currently no COS for studies of MCI. Collaboration with individuals with lived and professional experience throughout the process will support the development of a meaningful and useful COS to all stakeholders. The COS will help to standardise and encourage transparent outcome reporting in future MCI research and support collection of outcomes in clinical practice.

The study has been costed and designed with inclusivity and accessibility as a priority. For example, the study team have access to translation and interpreter services, interviews and surveys can be completed online or in person depending on participant preference and digital access, and the team have established links with community groups to encourage participation of underserved groups.

### Limitations

Initial searches identified a very high number of interventional studies in MCI and indicated that an umbrella review would be the most appropriate design for the literature search. By focusing only on studies which have been included in a systematic or scoping review, the review may not identify the most recently published studies. However, as the purpose of the review is to identify outcomes rather than intervention effectiveness, this is unlikely to affect the relevance of the review.

Study recruitment and data collection will predominantly be based at NHS sites in England and only articles available in English will be eligible for inclusion due to the importance of extracting outcomes verbatim. It is therefore possible that the generalisability of the findings may be limited to higher-income or Western countries. Efforts will be made to encourage a diverse and representative sample to contribute to the COS and outcomes identified in international literature will be included to mitigate this potential risk.

Finally, the study will identify the core outcomes for MCI and most common outcome measurement tools, but further research will be required to determine the recommended outcome measurement tools for the COS.

## Data Availability

Participants will be asked if they consent to data sharing beyond use in the COS development. Anonymised data may be shared upon reasonable request from the authors if the request is in line with consent received from participants.

## Ethics and dissemination

### Ethics approval/informed consent

The umbrella review is a secondary analysis of published work and does not require ethical review. Ethics approval for all other stages has been granted by London – Queen Square Research Ethics Committee (23/PR/1580). Valid informed consent will be received and documented ahead of eliciting survey responses or conducting interviews with prospective participants. Transcripts will be de-identified prior to analysis. All data will be reported in such a way that it is not possible to identify individual study participants.

### Dissemination

Dissemination is a fundamental part of the research process in developing a COS. The COS will be shared with key stakeholders (e.g., clinicians, researchers and research organisations, policymakers, funding agencies) and an accessible version will be developed for patients and the public. The COS will also be registered with the COMET database. The research findings from each stage of the study will be published in peer-reviewed academic journals and shared at international conference presentations and via social media and blog posts. Findings will be disseminated to participating stakeholders via a PPI co-produced Plain English report. We will work with key organisations (e.g., the Brain Health Clinic network and trial registries) to promote and facilitate wide adoption of the COS in research and relevant clinical settings^29^.

COS can help to improve the transparency of research and comparison and synthesis across multiple studies by encouraging reporting of a set of standardised outcomes.

Development and adoption of this COS is expected to enhance the value of interventional research into MCI and minimise research waste by encouraging reporting of agreed meaningful outcomes to all relevant stakeholders and most importantly outcomes which are important to patients with MCI.

## Acknowledgements

We would like to thank our team of PPI contributors and co-applicants for their insightful contributions towards study development and refinement.

## Author contributions

Study conception: NT, EC

Study design: NT, EC, VG, SH, AM, JC, JW, JD, WM, AR, SR, NW, SA

Manuscript authorship: VG, NT

Manuscript review: NT, EC

